# Implementation Determinants of Integrated TB Diabetes Care Package in Pakistan TB Control Program; A Mix Method Pragmatic Study Protocol

**DOI:** 10.1101/2025.06.30.25330571

**Authors:** Saima Aleem, Zohaib Khan, Saima Afaq

## Abstract

**Background:** Tuberculosis (TB) is a significant global health threat, particularly in high-burden countries like Pakistan. The incidence of diabetes mellitus among TB patients is rising, necessitating integrated clinical management to address the dual burden. Despite existing guidelines, the implementation of integrated TB diabetes care remains inadequate in Pakistan. This study aims to explore the implementation determinants of integrated TB diabetes care in the Pakistan TB Control Program.

**Methods:** This study aims to identify and assess the implementation determinants of an integrated TB-diabetes care package within Pakistan’s Provincial TB Control Program. A mixed-methods pragmatic approach guided by the Consolidated Framework for Implementation Research (CFIR) will be employed. The study focuses on both inner settings (organizational factors) and outer settings (external factors). Quantitative and qualitative data will be collected from stakeholders, including policy-makers, healthcare providers, and TB site staff.

**Analysis:** Systematic review will be guided by the Preferred Reporting Items for Systematic Reviews and Meta-Analyses (PRISMA-2020). Quantitative data will be analyzed using SPSS V.29 to assess organizational readiness for integrated care. The overall mean scores will interpret readiness levels, providing insights into the facilitators and barriers to implementation. Qualitative data will be analyzed using NVivo-12, following a framework analysis approach to identify key themes and categories. The study will quantify qualitative findings through a validated item rating scale, which is a 5-point Likert scale ranging from -2(Barrier) to 0 (neutral) to +2(Facilitator), to determine the valence and strength. The scores of -1 to +1 will identify the item as less barrier and facilitator, measuring the valence and strength of implementation determinants.

**Ethical Implications and Dissemination:** The study has received ethical approval from the Ethics Review Committee of Khyber Medical University. Study findings will be submitted for publication in peer-reviewed journals and will be presented at relevant conferences.

**Article Summary:** 

**Strength & Limitation of Study:**

- This study employs a pragmatic mixed-methods design, combining quantitative and qualitative approaches to identify implementation determinants for integrated tuberculosis and diabetes care packages in Pakistan.
- Barriers and facilitators to the implementation of TB-Diabetes integrated care will be identified across multiple TB centers including primary, secondary, and tertiary centers in five districts in two provinces.
- Patient perspective will not be investigated.

## INTRODUCTION

The dual burden of TB and DM has gained increasing attention globally over the past decade. In 2011, the World Health Organization and the International Union Against Tuberculosis and Lung Disease published a collaborative framework for Care and Control of Tuberculosis and Diabetes and recommended bidirectional screening and integrated management of TB and DM(1). Since then, several countries have piloted integrated care models with varying success levels.

Tuberculosis (TB) is a major global health threat, with an estimated 10.6 million people falling ill with tuberculosis (TB) worldwide, including 5.8 million men, 3.5 million women and 1.3 million children in 2022(2). Among the 30 countries with the highest TB burden, India has the highest incidence (26% of global cases) and Pakistan ranks 5th (5.8% of global cases). In Pakistan, TB prevalence is 348 per 100,000, incidence is 276 per 100,000, and mortality is 34 per 100,000(3)

Recent studies have revealed a growing link between TB and diabetes mellitus, particularly in South Asia. The prevalence of diabetes among patients with active TB has increased since 2000(4). Individuals with diabetes are more likely to develop TB, and diabetes often coexists with TB, requiring integrated clinical management(5). In Pakistan, inadequate diagnosis or management of diabetes in TB patients exacerbates health issues and strains healthcare resources, thereby affecting the economy(6).

The World Health Organization and the International Union Against Tuberculosis and Lung Disease advocate a collaborative approach for managing TB and diabetes. They recommend establishing mechanisms for collaboration between TB and diabetes control programs and for the detection and management of both conditions in each patient(7).

Bidirectional screening for TB and diabetes has been conducted in Asia(8, 9). However, challenges persist, including the inadequate integration of services and limited guideline implementation. In Indonesia, although guidelines for TB-DM co-management have been in place since 2015, their implementation has been hampered by financial constraints, insufficient resources, and weak coordination among healthcare facilities(10, 11)

In Pakistan, TB control is managed through National and Provincial TB Control Programs. Despite their efficiency, integrating diabetes screening and management with TB control remains inadequate(12). China and India, which bear the highest global burdens of both tuberculosis (TB) and diabetes mellitus (DM), are leading the implementation of integrated screening and care models. In China, a large-scale study conducted across multiple provinces demonstrated the feasibility of screening DM patients for active TB, revealing a TB prevalence of 0.80%(13). This study also highlighted several operational challenges, including insufficient human resources and the need for more streamlined screening algorithms. In India, pilot studies conducted in diverse settings have yielded promising outcomes while also identifying context-specific obstacles. For instance, a study in Kerala found that integrating DM screening into Directly Observed Treatment, Short-Course (DOTS) clinics was feasible and uncovered a high DM prevalence of 44% among TB patients(14).

Beyond South Asia, relevant perceptions have been drawn from experiences in sub-Saharan Africa. A mixed-methods study in Ethiopia identified critical challenges such as poor coordination between TB and DM programs, inadequate diagnostic infrastructure, and low awareness among healthcare workers(15). These findings emphasize the need to strengthen health systems to support the integration of TB and DM care.

Further implementation studies in other low- and middle-income countries have identified similar challenges. Studies conducted in India reported significant barriers to integration, including a lack of awareness among healthcare providers, inadequate infrastructure, and poor coordination between TB and DM programs(16, 17). Similarly, Workneh et al. also stressed upon the importance of health system readiness, including staff training and effective supply chain management, for the successful implementation of integrated care(15).

In designing integrated TB-DM care programs, it is imperative to address the facilitating and hindering factors both within and outside the healthcare system. Previous implementation studies report that inner setting factors, particularly structural characteristics, organizational readiness, implementation culture and climate significantly influence the success of integrated TB-diabetes care implementation(18). Additionally, the readiness for implementation will vary across different levels of the TB healthcare system. Lastly, evidence also highlights that outer setting factors including cosmopolitanism, peer pressure, and external policies play a critical role in facilitating or hindering the implementation process(19, 20).

Pakistan’s healthcare system, characterized by its complexity and hierarchical nature, presents significant challenges for the integration of new innovations. Although prior studies have focused on the screening of comorbid diabetes among TB patients, the implementation of integrated management strategies within the TB control program remains limited. Notably, many of the challenges associated with implementation have been identified during the post-implementation phase, rather than being anticipated and addressed in earlier stages. Although the guidelines for integrated TB-diabetes care are available, there is a significant gap in their implementation within Pakistan’s healthcare system. The perspectives of frontline health workers and program managers regarding implementation challenges are also underexplored. By identifying key barriers and facilitators, the strategies to improve the integration of diabetes screening and management within the existing TB control infrastructure and its effective implementation can be designed.

### Study Rationale

Given Pakistan’s high dual burden of TB and DM, there is an urgent need to identify contextually appropriate strategies for scaling up the integrated services nationwide. For this reason, this study aimed to identify the internal and external determinants to implement the Tuberculosis Diabetes Care Package in the Provincial TB Control Program at pre and during implementation. The integrated TB-Diabetes care package will be designed, implemented, and evaluated for screening, preventing, and managing diabetes in people with TB at the primary, secondary, and tertiary care TB facilities under the Grand Challenge Fund project (Ref # 20-GCF-770/RGM/R&ID/HEC/2021) in Pakistan & supported by the World Bank and implemented by the Higher Education Commission (HEC).

This current study aims to address the key evidence gaps by (i) exploring health system barriers and facilitators for TB-DM integrated care implementation across diverse TB settings in Khyber Pakhtunkhwa (KPK), Pakistan and (ii) eliciting the perspectives of multiple stakeholders, including frontline providers and program managers, policymakers and financial stakeholders. By generating rich quantitative and qualitative insights into barriers and facilitators to the implementation of integrated care, this study will inform policy and practice to strengthen TB-DM collaborative activities in Pakistan and similar high-burden settings.

## RESEARCH QUESTION

What are the implementation determinants of the integrated TB diabetes care package in the Pakistan TB Control Program?

## OBJECTIVE(S)

Objective of this study is to: (i) review the implementation determinants of TB DM integrated care in South Asian Association of Regional Corporation (SAARC) (ii) explore the implementation determinants of the integrated TB Diabetes care external and internal to the Provincial TB control programs in Pakistan (iii) systematically evaluate variations in the implementation strategies of the integrated TB and diabetes care package across healthcare facilities using a structured evaluation framework and to analyze the relationship between these strategies and key evaluation outcomes and (iv) assess which determinant will contribute both in valence (positive or negative influence) and extent (combined influence) to the TBDM integrated care package implementation.

## METHODS

This mixed methods pragmatic study will be guided by the Consolidated Framework for Implementation Research (CFIR). The “Consolidated Framework for Implementation Research” (CFIR), published in 2009, includes five domains and 39 constructs that facilitate the identification of implementation determinants, allowing selection based on the intervention’s context and stage (Figure 1). Two CFIR domains are particularly relevant: inner and outer settings. The inner settings domain addresses organizational factors, such as structural characteristics, culture, and readiness for implementation (Figure 1). The outer settings domain considers external factors such as policies, peer pressure, and cosmopolitanism, which influence an organization’s ability to implement interventions effectively(21).

**Figure 1:**
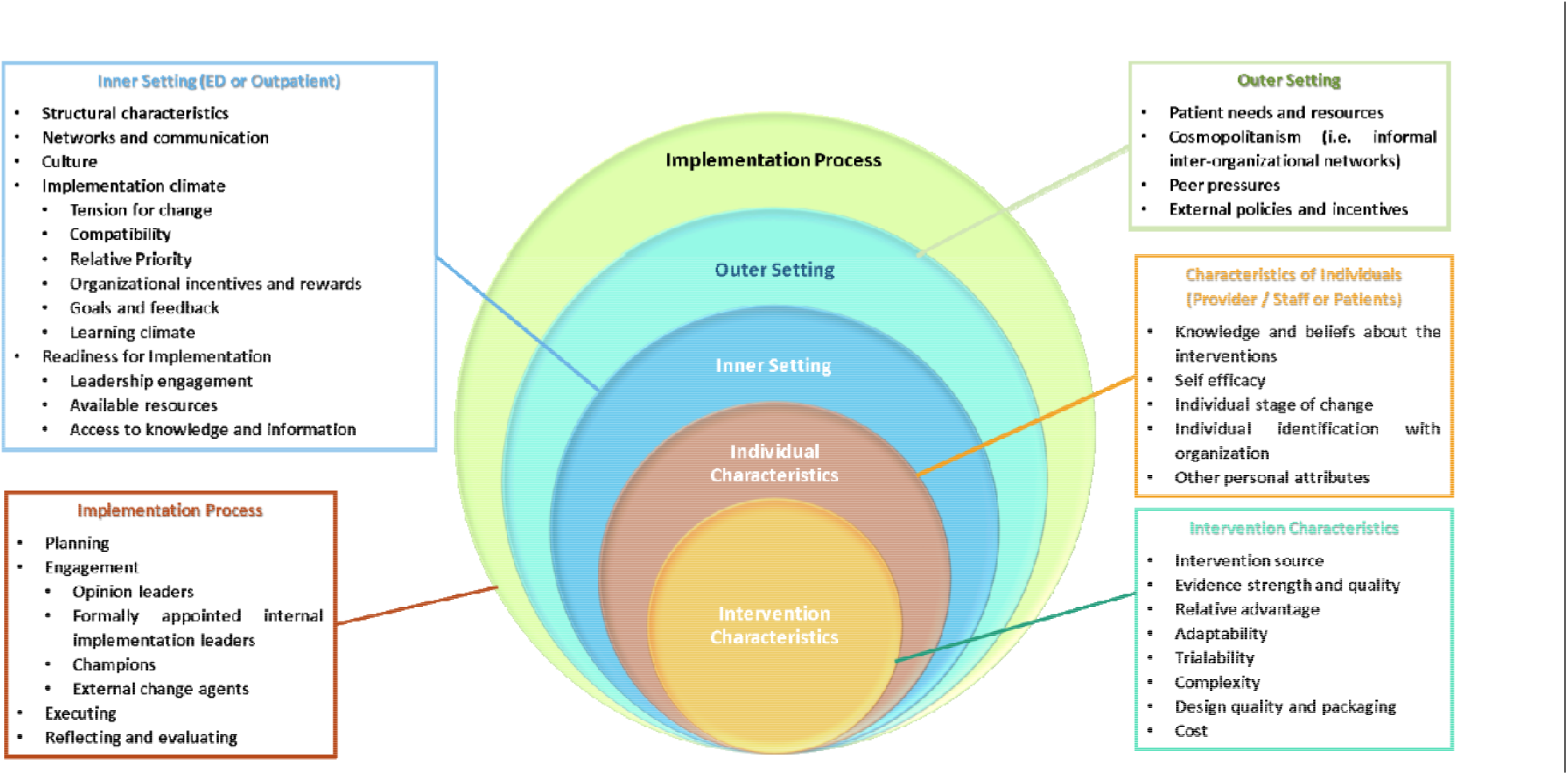
CFIR Domains & Constructs.

## THEORETICAL FRAMEWORK

### Objective 1

#### Systematic Review

The dual burden of tuberculosis (TB) and diabetes mellitus (DM) poses a significant public health challenge, particularly in countries of the South Asian Association for Regional Cooperation (SAARC). The bidirectional relationship between TB and DM not only increases disease burden but also complicates diagnosis, management, and outcomes. While the World Health Organization (WHO) has called for integrated care models, the implementation of such strategies in real-world health systems remains inconsistent. There is currently no synthesis of how integrated TB-DM care is being operationalized across the SAARC region or what factors influence its success. This systematic review aims to identify and analyze implementation determinants of integrated TB and DM care in SAARC countries.

This review will include qualitative, quantitative, and mixed-methods primary studies reporting on the implementation of integrated TB-DM care in SAARC countries. A comprehensive search will be conducted in MEDLINE (via Ovid), EMBASE, Web of Science, Cochrane CENTRAL, and CINAHL. Grey literature will be sourced from Open Access Theses and Dissertations (OATD) and Google Scholar. Two reviewers will independently screen title and abstract and full text using Rayyan and using a structured Excel form, three reviewers will extract data. Quality assessment will be done by Quality assessment was conducted by using Mixed Methods Appraisal Tool (MMAT), Critical Appraisal Skills Programme (CASP) (for qualitative studies) and Joanna Briggs Institute (JBI) critical appraisal tools (for quantitative studies). A narrative synthesis will be conducted in line with SWiM guidelines to categorize implementation determinants as barriers or facilitators, and to explore patterns across countries and contexts.

This will be the first systematic review to synthesize evidence on the implementation of integrated TB and DM care in the SAARC region. The findings will inform policymakers, program managers, and regional stakeholders by highlighting context-specific challenges and enablers of integration. The review will also identify gaps in current literature and provide direction for future research and program design. The findings from the systematic review will also guide the qualitative and quantitative studies for objective 2

### Objective 2

#### Study Settings

- To identify the implementation determinants to provincial TB facilities, settings will include organizations external to and having direct relations to inner settings, including the Global Fund, National TB Control Program Pakistan, Ministry of National Health Services, Regulations and Coordination Pakistan, Provincial TB Control Program (KPK and Punjab), and provincial health departments.
- The inner settings will include 13 TB health facilities in four districts in KPK and one district of Punjab where an integrated care package for TBDM will be implemented.

### Sample Size and Sampling Technique

For outer settings, based on purposive sampling, policymakers, and stakeholders mentioned in Table 1 will be included.

**Table 1:**
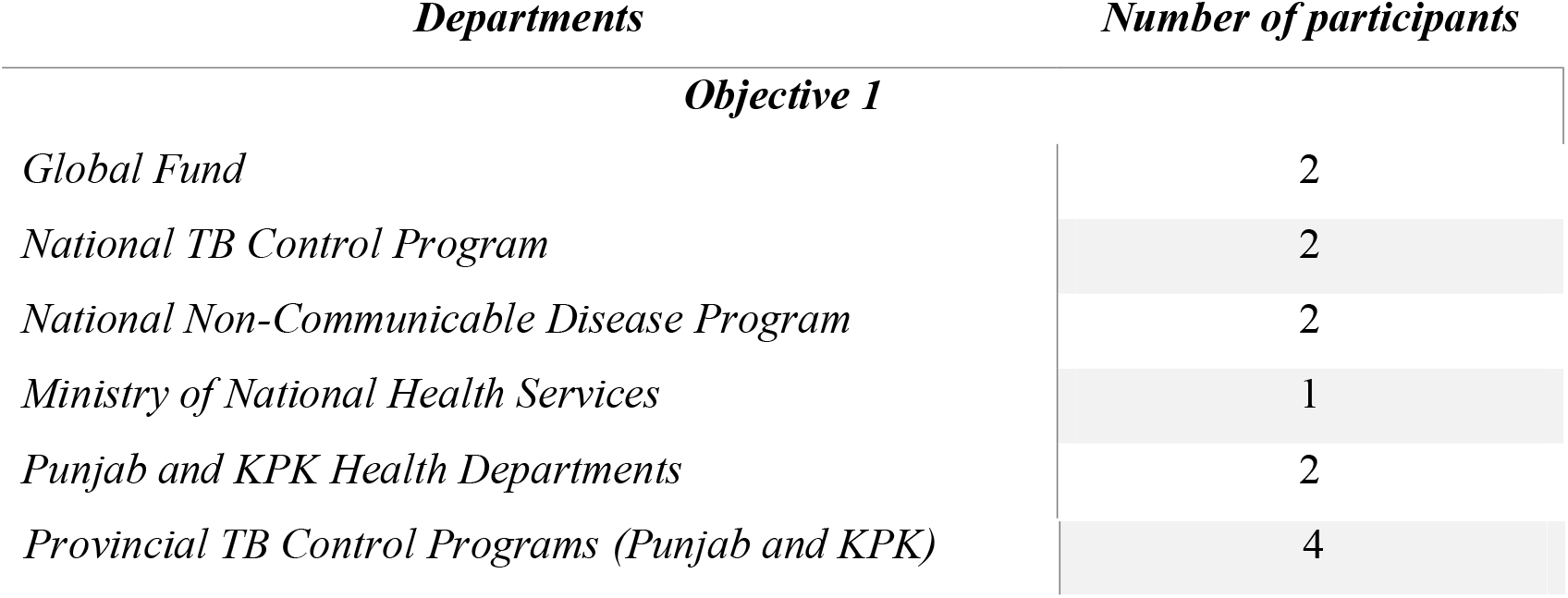
Number of participants for qualitative assessment of outer setting.

For the outer settings, key stakeholders who have in-depth knowledge and influence over the TB-diabetes policy and implementation at the national and provincial levels will be purposively selected. This approach will ensure that we capture diverse perspectives from decision makers and program managers who are directly involved in shaping integrated care strategies. To minimize and mitigate the selection bias, we strive for maximum variation in our purposive sample, ensuring representation at different organizational levels and geographic areas. Additionally, we will carefully document our sampling decisions and consider their potential impact during data analysis and interpretation.

For inner settings, by using census sampling, we will select all the staff from the 13 TB facilities in Khyber Pakhtunkhwa from Peshawar, Swat, Mardan and Abbottabad and from Rawalpindi in Punjab.

Use of census sampling of all staff from the selected TB health facilities will ensure comprehensive coverage by frontline implementers. This approach allows us to capture the full range of experiences and perspectives within each facility, providing a holistic view of the implementation determinants at the operational or activity level.

## DATA COLLECTION

Informed consent will be obtained from all study participants. Participants will be informed about the components of the study and the importance of their responses before data collection. Confidentiality of all study participants was ensured.

### For Outer Settings

In-depth interviews will be conducted with all study participants using the topic guides adopted from the CFIR Outer Setting Qualitative Guide covering seven constructs and three sub-constructs (Figure 2)

**Figure 2:**
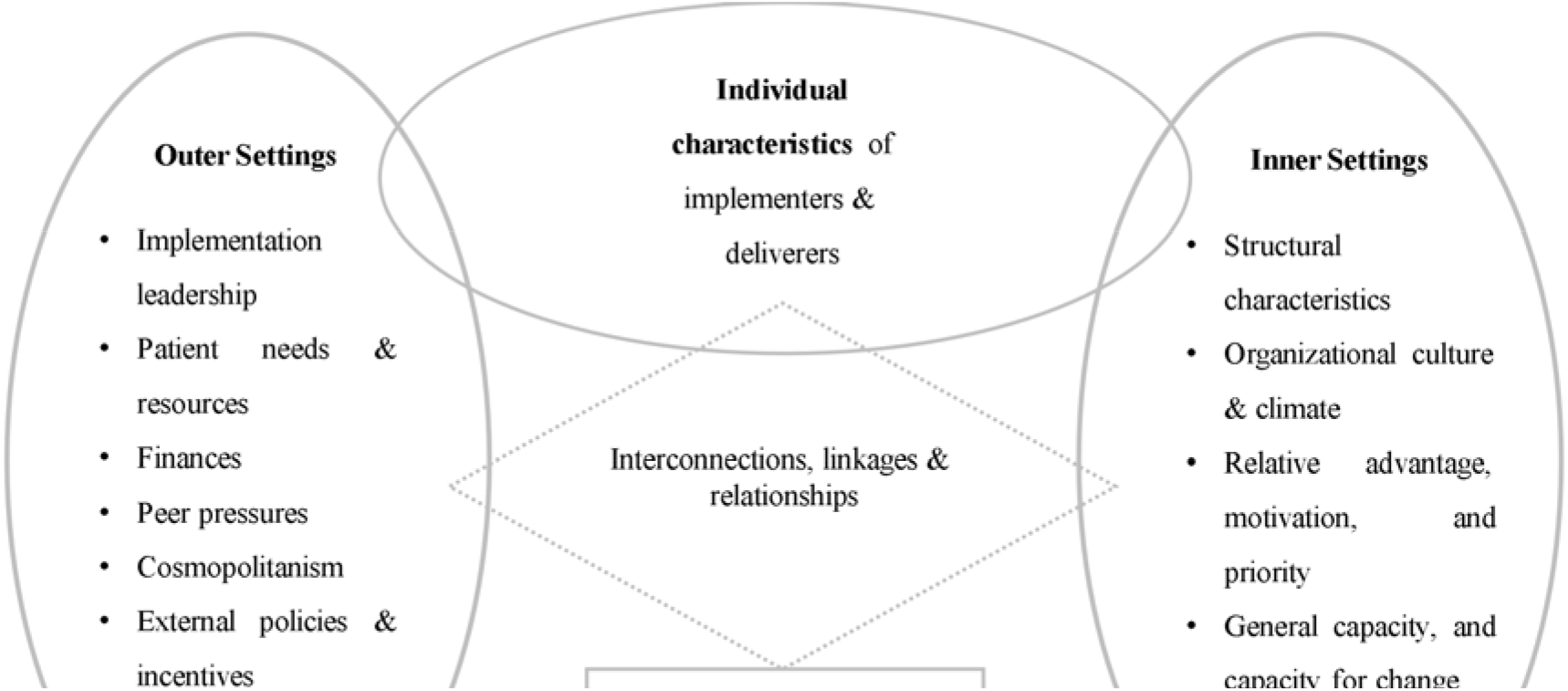
Intersectionality for Implementation Determinants.

### For Inner Settings

At TB health facilities, structural characteristics, including physical infrastructure, information technology infrastructure, and work infrastructure, availability of diabetes services and readiness for integrated care will be assessed using a checklist adapted from the World Health Organization Availability and Readiness Assessment tool.

For the organizational readiness to integrated care assessment, based on **R= MC**^**2**^ which is a readiness heuristic reflecting implementation barriers and facilitators across three components: motivation, general capacity, and capacity for change(22), from the published literature, a 5-point Likert scale-based data collection tool will be adapted and contextualized. After the quantitative assessment, in-depth interviews will be conducted with all study participants in inner and outer settings using the topic guides adapted from the CFIR qualitative guide (Figure 3).

**Figure 3:**
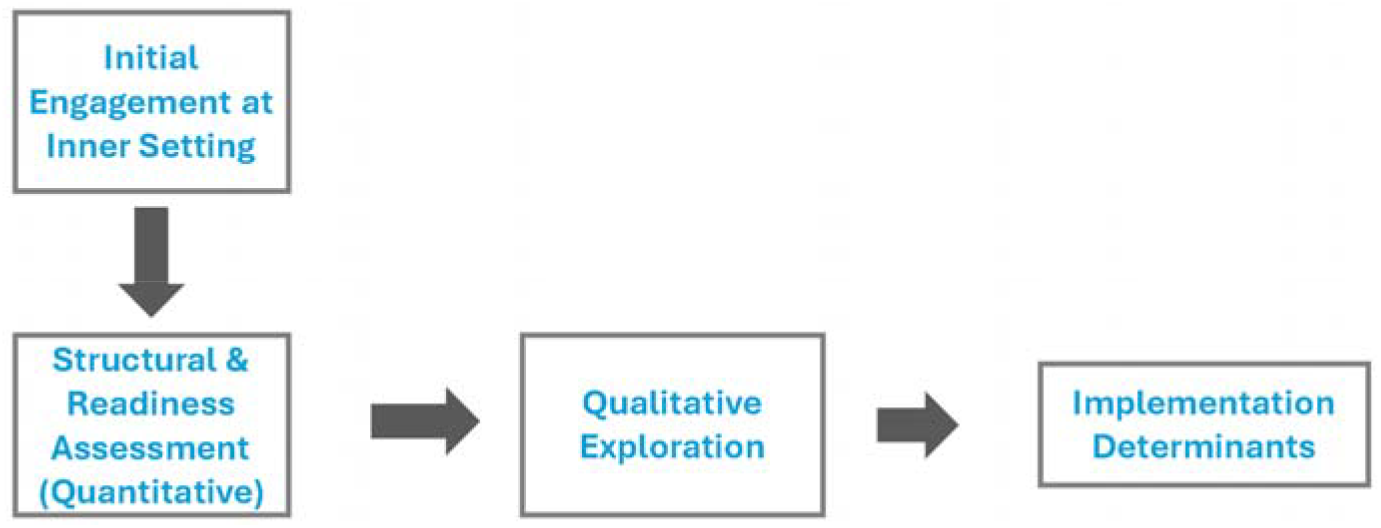
Inner Setting Methodology.

The adaptation process will involve the following steps:

1. Review of the original CFIR guides.
2. Modification of questions to reflect the specific context of TB-diabetes integrated care in Pakistan.
3. A small-scale pilot testing will be performed with a small sample of participants, preferably two for each guide.
4. Refinement to the final topic guides will be made based on pilot test feedback.

This process will ensure that the interview guides are culturally appropriate, comprehensible, and effective in eliciting relevant information about implementation determinants.

After the interview guide pilot study, definitive interviews will be conducted with all the study participants.

## DATA ANALYSIS PROCEDURE

### Integration of Quantitative and Qualitative Data

To leverage the strengths of our mixed-methods approach, we will integrate quantitative and qualitative data using a convergent parallel design. Integration will occur at multiple levels. We will conduct quantitative assessments of structural readiness and readiness for integrated care, which will guide our qualitative interviews to explore the identified issues more deeply. The analysis will utilize a joint display approach to visually represent the quantification of qualitative data. For interpretation, we will integrate findings from both quantitative and qualitative strands using a weaving approach, presenting the results together on a theme-by-theme basis for each major implementation determinant identified.

### Qualitative Analysis

All interviews (IDIs) will be audio-recorded and transcribed verbatim. The transcripts will be analyzed using NVivo-12. The data obtained from the interviews conducted using the Inner-Outer Setting Domain of the CFIR Model will be analyzed inductively and deductively using a framework analysis approach.

The researcher SA will familiarize herself with all the data, and the categories of the framework will be developed deductively based on a CFIR model and inductively developed from the data of preliminary interviews. Familiarization and framework development will be followed by further steps of indexing, charting, and interpretation of data using MS Excel for the matrix.

### Quantitative Analysis

Quantitative analysis for the structural readiness and readiness for integrated TB Diabetes care will be conducted using SPSS V. 26. Readiness for integrated care will be scored by rating each item in a section on a scale of 1 to 5. We will calculate the mean score for each section by summing the scores of all items and dividing them by the number of items. To obtain the overall mean score, we will sum the mean scores of all sections and divide by the number of sections. Finally, we will interpret the overall mean score using predefined ranges to determine the readiness level.

### Section Mean Score Calculation

For each section (Motivation, Capacity, Culture, Barriers and Facilitators etc), the mean score will be calculated as follows:

Mean Score for Section=∑Item Scores/Number of Items in Section

### Overall Mean Score Calculation

Overall Mean Score=∑Mean Scores of All Sections/Number of Sections. Table 2 presents the criteria for the interpretation of the scores.

**Table 2:**
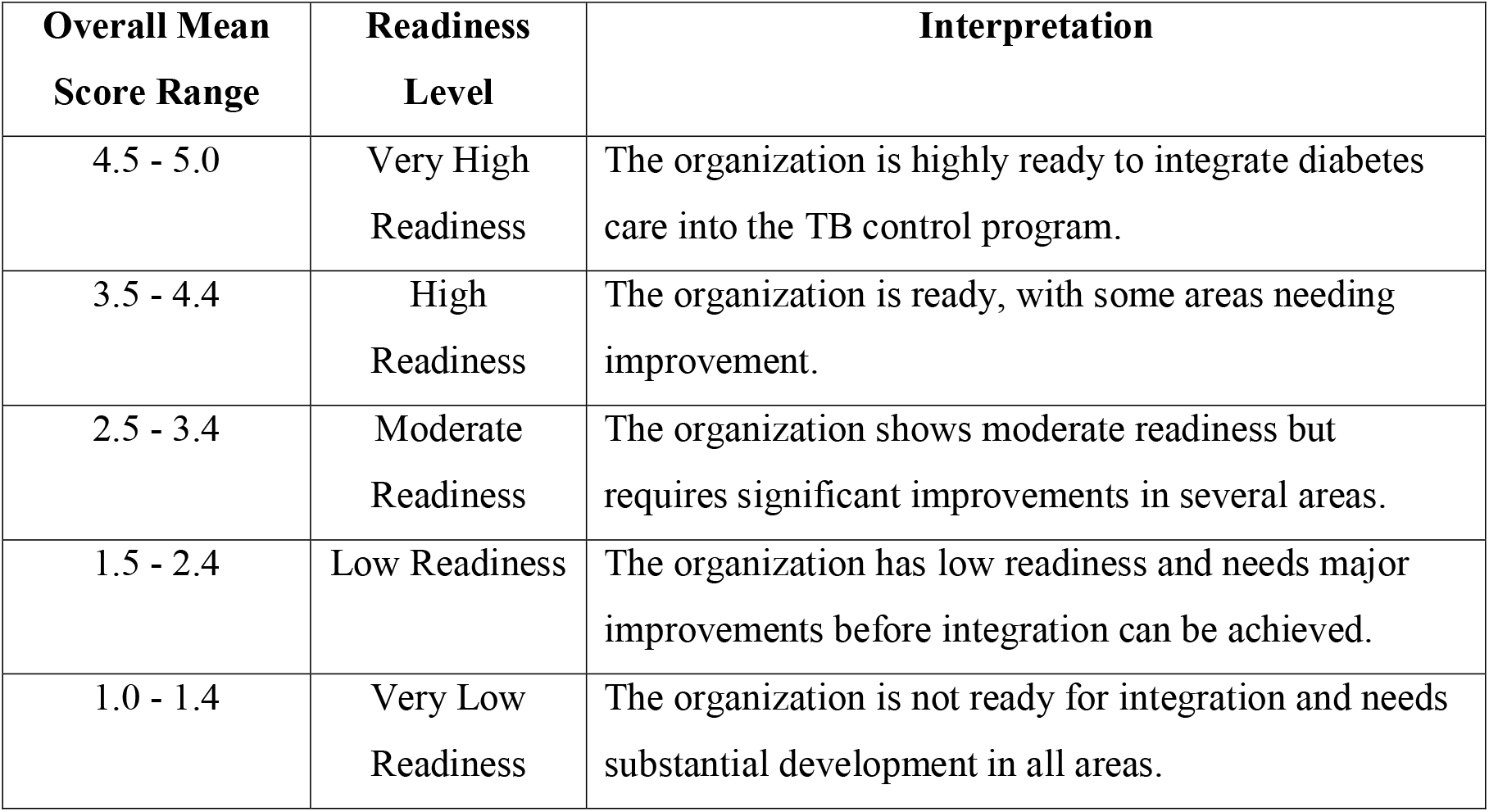
Interpretation of Scores.

To highlight variations in readiness, a heat table coding system will be applied to identify specific areas of strengths, weaknesses, and trends(23).

### Objective 3

To evaluate the variation across the implementation strategies, a comprehensive study will be conducted across 13 healthcare facilities in Pakistan as presented in figure 4. The study will begin with the development of a scoring system following a detailed literature review of existing implementation frameworks and scoring methodologies to identify best practices. Key domains will include but not limited to Governance & Coordination, Screening & Case

**Figure 4:**
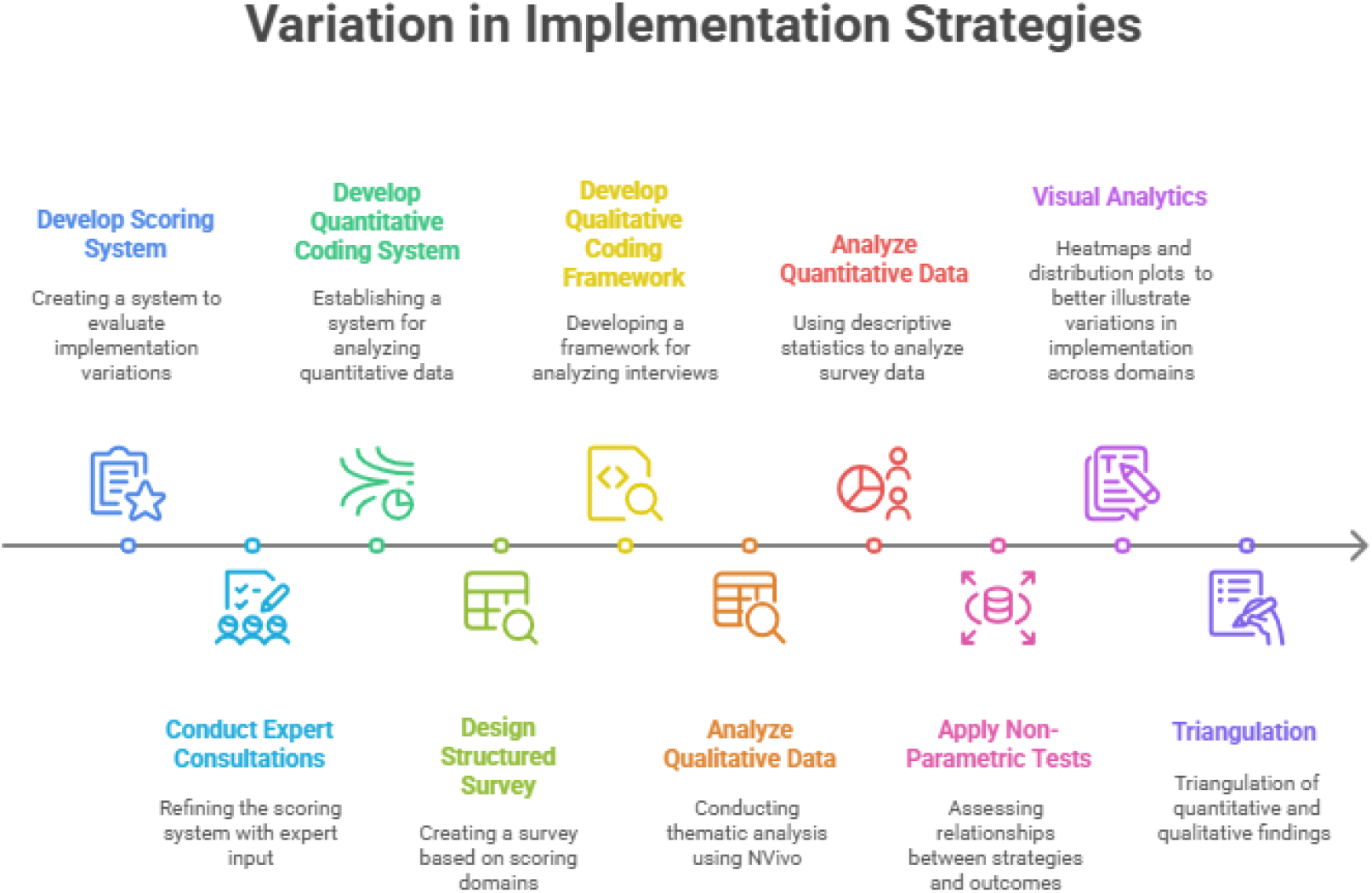
Roadmap for Assessment of Variation in Implementation Strategies.

Detection, Treatment & Clinical Outcomes, Capacity Building & Workforce Engagement, Patient & Community Engagement, and Sustainability & Scalability. A preliminary scoring rubric will be created to reflect the continuum from no implementation to full implementation, and refined through expert consultations. Subsequently, structured surveys will be designed based on the identified scoring domains to support quantitative data collection.

Simultaneously, a qualitative coding framework will be developed to guide the thematic analysis of interviews with facility managers, healthcare providers, and policymakers, focusing on barriers, facilitators, and implementation variations. Once data collection is complete, qualitative data will be transcribed and analyzed thematically using NVivo, with systematic coding to identify major themes such as common implementation barriers and effective strategies. Quantitative data will be analyzed using descriptive statistics to evaluate implementation scores across facilities, and a correlation matrix will be developed to explore patterns in the relationships between implementation strategies and key outcomes. This integrated dataset will serve as the foundation for further statistical testing.

To assess these relationships rigorously, non-parametric statistical tests, including Spearman’s correlation will be applied, to measure the strength of associations between implementation strategies and performance outcomes. Visual tools such as heatmaps and distribution plots will be used to illustrate domain-specific variations. Triangulation of qualitative and quantitative findings will offer a comprehensive understanding of the implementation process and its effectiveness.

### Objective 4

#### Quantification of Qualitative Data

To understand which determinant will contribute to both in valence (positive or negative influence) and extent (combined influence) to the TBDM integrated care package implementation, quantitative analysis of qualitative data will be conducted using the validated item rating scale, which is a 5-point Likert scale ranging from -2(barrier) to 0 (neutral) to +2(facilitator), to determine valence and strength. Scores of -1 to +1 identify the item as having fewer barriers and facilitators.

## Data Availability

The principal investigator, SAL, will be primarily responsible for publishing the study results according to the study's publication and dissemination plan and all the findings will contribute to her doctoral dissertation. The policy brief which will be developed based on all the findings will be shared by the Provincial TB Control Programs, National TB Control Program and all relavant stakeholder at provincial, national and international level to strengthen TB DM integrated care.

## ETHICS AND DISSEMINATION

### Research ethics approval

This study was approved by the Ethics Review Committee (ERC) of the Institute of Public Health & Social Sciences, Khyber Medical University, Peshawar (Ref # KMU/IPHSS/Ethics/2024/EG/200).

### Consent

The lead researcher will contact study participants via telephone or in-person to invite them to participate in a semi-structured interview at the time and location of their choice, including online options. Before conducting the interview, the researcher will explain the study and its purpose, providing participants with an information sheet and consent form. Participants will be encouraged to ask questions or express concerns and will be invited to sign the consent form.

Before beginning the interview, the researcher will inform participants that they may stop the interview at any time and withdraw their consent to participate, either during or after the interview. If a participant withdraws, their interview recordings and transcripts will be excluded from the study and destroyed. Withdrawal from the study will not affect participants’ employment or roles in any way.

When contacting participants to schedule an interview, the researcher will explain the purpose of the follow-up interview. Participants will have the opportunity to ask questions or express concerns and will be invited to sign a consent form. Before starting the interview, the researcher will reiterate that participants can stop the interview at any time and withdraw their consent, with their data being removed and destroyed upon withdrawal.

### Confidentiality

The confidentiality of the participants will be rigorously maintained by the researcher. The study protocols, documentation, data, and all other generated information will be kept strictly confidential and stored in password-protected electronic files. No study-related information or data will be released to unauthorized third parties without prior written approval from a researcher or supervisor.

### Implications for Policy and Practice

The findings of this study will have the potential to directly influence policies and practices within TB control programs in Pakistan.

1. The identification of key implementation determinants will allow policymakers to prioritize interventions that address the most critical barriers and leverage existing facilitators.
2. The readiness assessment results will inform targeted capacity-building efforts, ensuring that resources are efficiently allocated to areas of greatest need.
3. Insights from frontline implementers can guide the development of practical context-specific strategies for effective implementation of diabetes care into the existing TB control infrastructure.
4. The findings of this study can inform the development of policy brief, revision or development of national guidelines for integrated TB-diabetes care, ensuring that they are grounded in local realities and implementation principles.

### Competing interests

None

### Patient and public involvement

Considering the nature of the study, the patients and/or the public were not involved in the design of this research.

### Data statement

Once the study is completed, the data will be available from the corresponding author on request. Systematic review, quantitative and qualitative data generated or analyzed during the study will be included in manuscripts to be submitted for publication in peer-reviewed journals.

### Funding

This Ph.D. research of Dr. Saima Aleem is supported by the Higher Education Commission Pakistan (Reference# 20-GCF-770/RGM/R&ID/HEC/2021).

### Authors Contributions

SAL and SA* conceived the research concept. SAL drafted the research protocol. ZK contributed to the refinement of the research methodology. All authors read, provided important revisions, and approved the final version of the manuscript.

## Notes

### Competing Interest Statement

The authors have declared no competing interest.

### Funding Statement

The Ph.D. research of Dr. Saima Aleem is supported by the Higher Education Commission Pakistan (Reference# 20-GCF-770/RGM/R&ID/HEC/2021).

